# Post-stroke fatigue is linked to resting state posterior hypoactivity and prefrontal hyperactivity

**DOI:** 10.1101/2021.03.17.21253777

**Authors:** Georgia Cotter, Mohamed Salah Khlif, Laura Bird, Mark E Howard, Amy Brodtmann, Natalia Egorova

## Abstract

**Background and Purpose:** Fatigue is associated with poor functional outcomes and increased mortality following stroke. Survivors identify fatigue as one of their key unmet needs. Despite the growing body of research into post-stroke fatigue, the specific neural mechanisms remain largely unknown.

**Methods:** This observational study included 63 stroke survivors (22 women; age 30-89 years; mean 67.5±13.4 years) from the Cognition And Neocortical Volume After Stroke (CANVAS) study, a cohort study examining cognition, mood, and brain volume in stroke survivors following ischaemic stroke. Participants underwent brain imaging 3 months post-stroke, including a 7-minute resting state fMRI echoplanar sequence. We calculated the fractional amplitude of low-frequency fluctuations, a measure of resting state brain activity at the whole-brain level.

**Results:** Forty-five participants reported experiencing post-stroke fatigue as measured by an item on the Patient Health Questionnaire-9. A generalised linear regression model analysis with age, sex, and stroke severity covariates was conducted to compare resting state brain activity in the 0.01-0.08 Hz range, as well as its subcomponents - slow-5 (0.01-0.027 Hz), and slow-4 (0.027-0.073 Hz) frequency bands between fatigued and non-fatigued participants. We found no significant associations between post-stroke fatigue and ischaemic stroke lesion location or stroke volume. However, in the overall 0.01-0.08 Hz band, participants with post-stroke fatigue demonstrated significantly lower resting-state activity in the calcarine cortex (p<0.001, cluster-corrected p_FDR_=0.009, k=63) and lingual gyrus (p<0.001, cluster-corrected p_FDR_=0.025, k=42) and significantly higher activity in the medial prefrontal cortex (p<0.001, cluster-corrected p_FDR_=0.03, k=45), attributed to slow-4 and slow-5 oscillations, respectively.

**Conclusions:** Post-stroke fatigue is associated with posterior hypoactivity and prefrontal hyperactivity, reflecting dysfunction within large-scale brain systems such as fronto-striatal-thalamic and frontal-occipital networks. These systems in turn might reflect a relationship between post-stroke fatigue and abnormalities in executive and visual functioning. This first whole-brain resting-state study provides new targets for further investigation of post-stroke fatigue beyond the lesion approach.

## Introduction

Post-stroke fatigue (PSF) is a common sequela of ischaemic or haemorrhagic stroke and is reported by stroke survivors as one of their most disruptive symptoms^1, 2^. It is associated with poor functional outcomes and reduced quality of life (QoL), a reduction in the likelihood of returning to work, and increased mortality.^1, 3, 4^ Prevalence rates of PSF vary from 29% to 92% of stroke survivors,^5-7^ far exceeding the 10-23% prevalence of fatigue in the general population.^8-11^

Prior studies aiming to identify the neural mechanisms of PSF have focused on associations with stroke types and lesion locations.^12^ No associations have been found with either ischaemic or haemorrhagic stroke,^13, 14^ nor stroke severity, stroke side, or infarct volume.^15^ In contrast, stroke type (e.g., lacunar, partial or total anterior circulation, posterior circulation) has been found predictive of PSF.^16^ Posterior circulation infarctions (POCIs)^16^ which account for roughly 20% of ischaemic strokes,^17^ have been linked to higher levels of fatigue than other stroke subtypes.^18-20^

Focusing on anatomical lesion locations, many studies did not find any links with PSF.^21-23^ Some authors related PSF to right-sided lesions,^24^ infratentorial region,^25^ caudate and putamen,^26^ right insula and anterior cingulate cortex,^27^ white matter lesions,^28^ and vertebrobasilar arterial infarcts through interaction with the modified Rankin Scale.^19^ Despite much heterogeneity, PSF has been associated with lesions in the posterior circulation territory, especially structures supplied by the vertebrobasilar arterial system including the brainstem, cerebellum, midbrain, and thalamus,^29-32^ reinforcing the association between PSF and POCI.

To date, few researchers have utilised functional MRI to explore the neural correlates of PSF. An association between high fatigue and perceived effort in stroke and a greater activation in the pre-supplementary motor area and the ipsilateral inferior frontal gyrus has been reported,^33^ as well as functional connectivity in the fronto-striato-thalamic (FST) network^34^ predictive of the response to the alertness-promoting treatment modafinil for PSF.^35^ However, the FST circuitry has been implicated in fatigue in other conditions, including in multiple sclerosis,^36^ granulomatosis with polyangiitis,^37^ traumatic brain injury,^38, 39^ Parkinson’s disease,^40^ and several non-neurological diseases such as HIV infection,^41^ ankylosing spondylitis,^42^ and chronic fatigue syndrome (CFS).^43^

We aimed to identify the neural correlates of PSF in terms of stroke type, lesion locations, and resting-state brain functioning at the whole-brain level. Specifically, we measured the fractional amplitude of low-frequency fluctuations (fALFF), computed as the relative contribution of the low-frequency band (0.01 – 0.08 Hz) to the entire detectable frequency range (0 – 0.25 Hz). fALFF indexes local spontaneous neuronal activity^44^ and thus is suitable to probe the neural basis of PSF. Furthermore, as specific frequency bands may differentially contribute to resting-state activity,^45^ we examined the slow-5 (0.01 – 0.027 Hz) and slow-4 (0.027 – 0.073 Hz) bands which are reported to reflect grey matter signals.^45, 46^ We hypothesised that there would be an association between PSF and posterior circulation ischaemic stroke, posterior lesion locations, as well as a difference in the resting-state neural activity between stroke survivors with and without PSF in the FST network.

## Materials and methods

### Participants

Ischaemic stroke patients from the Stroke Units at Austin Hospital, Box Hill Hospital, and the Royal Melbourne Hospital in Melbourne, Australia, were recruited as part of the Cognition And Neocortical Volume After Stroke (CANVAS) study.^47^ Ethical approval for the CANVAS study was obtained from each hospital’s Human Research Ethics Committee according to the Declaration of Helsinki and all participants provided written informed consent. Additional approval for the current study was obtained from the University of Melbourne’s Psychological Sciences Human Ethics Advisory Group.

All participants engaged in a structured interview to obtain information regarding medical history, current medications, and vascular risk factors. At 3 months post-stroke, participants completed MRI scanning, mood scales, and cognitive testing. Participants with both imaging (structural and functional images) and behavioural data at 3 months post-stroke were included in the analysis. Participants were excluded if they were unable to give informed consent, had experienced a haemorrhagic stroke or venous infarction, or had significant medical comorbidities making it unlikely that they would survive 3 years. Additional exclusion criteria included significant psychiatric history prior to stroke, or MRI ineligibility (e.g., claustrophobia or safety contraindications).

### Fatigue assessment

All participants completed the Patient Health Questionnaire-9 (PHQ-9)^48^: a self-administered screening measure of depression symptom severity. Participants rated each of the 9 symptoms of depression from the Diagnostic and Statistical Manual of Mental Disorders (DSM-V)^49^ on a 4-point scale of symptom frequency over the prior two weeks (0 = *not at all*, 1 = *several days*, 2 = *more than half the days*, or 3 = *nearly every day*). Fatigue was assessed via item 4 of the PHQ-9, “feeling tired or having little energy”, which shows good correspondence to the Fatigue Questionnaire items 1 (tiredness), 2 (need more rest), 3 (feels sleepy and drowsy), and 5 (lacking energy) as well as the total fatigue score, with correlations ranging between 0.52 and 0.61.^50^ We interpreted a score > 0 as indicating the presence of PSF.

### Assessment of stroke characteristics, cognitive function, and medical comorbidities

Strokes were classified in terms of stroke aetiology^51^ and stroke subtype^16^ by a stroke neurologist. The National Institutes of Health Stroke Scale (NIHSS)^52^ and the Modified Rankin Scale (mRS)^53^ were used to assess participant stroke impairment and disability. The presence of depression at 3 months post-stroke was determined by a global PHQ-9 score ≥ 5 and anxiety was assessed with the Generalised Anxiety Disorder questionnaire (GAD-7)^54^. Participants were screened for vascular and stroke risk factors including type 2 diabetes mellitus (T2DM), ischaemic heart disease, atrial fibrillation, and hypertension. Cognitive profiles of participants were assessed and reported in Table 1.

**Table 1.**
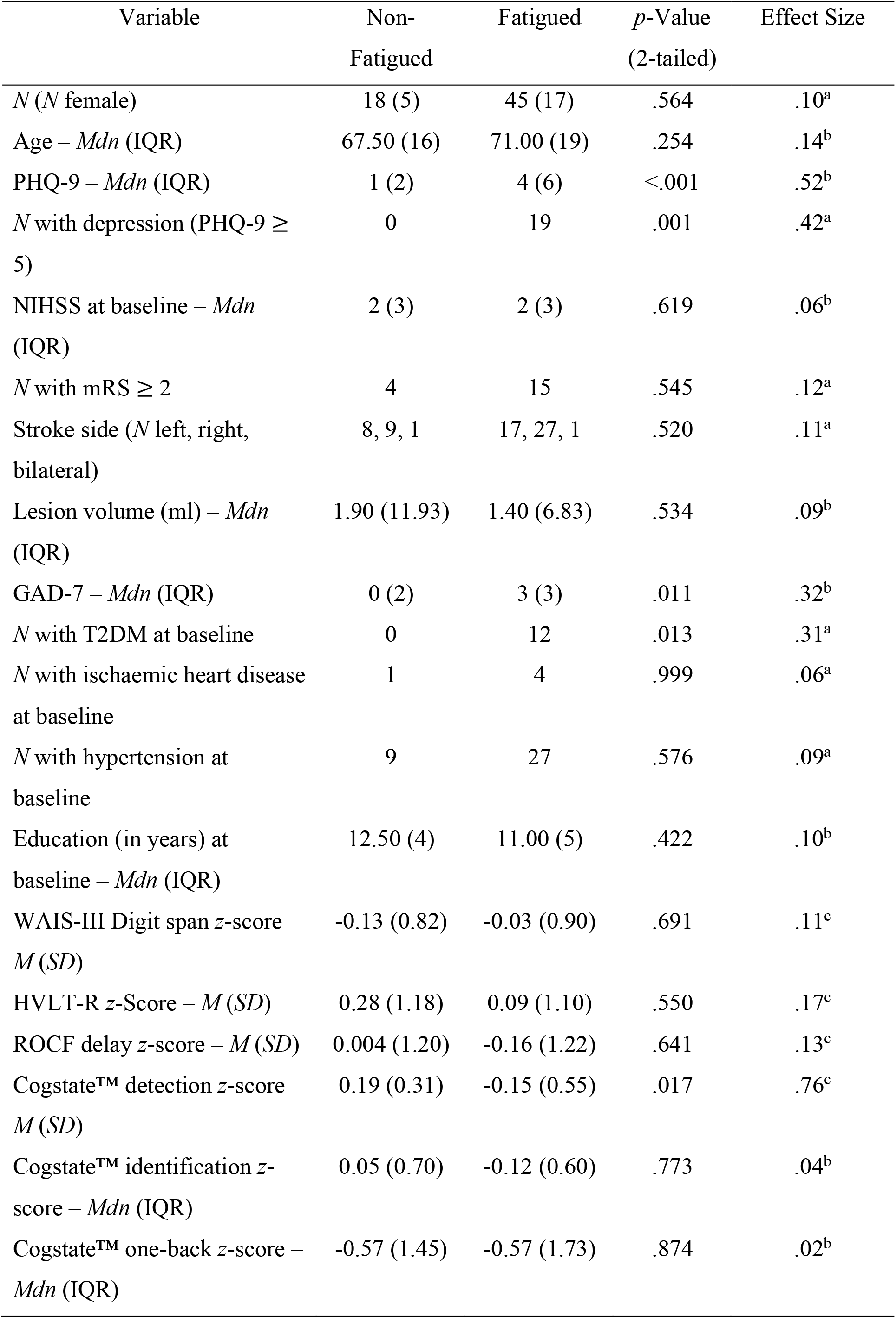
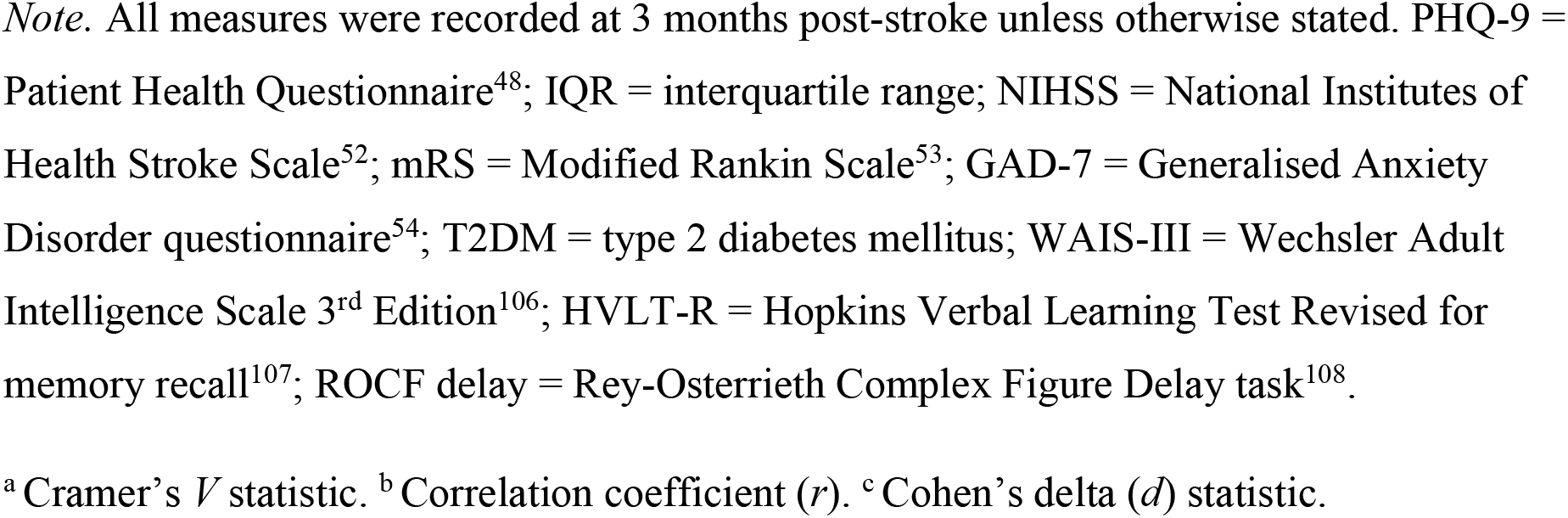
Group Comparison of Demographics and Clinical Outcomes. p-values are for the independent-samples t-test (digit span z-score, HVLT-R total learning z-score, ROCF Delay z-score, Cogstate™ detection z-score), a Mann-Whitney U test (age, PHQ-9 score, NIHSS at baseline, GAD-7 score, years of education at baseline, Cogstate™ identification z-score, Cogstate™ one-back z-score, lesion volume), a Pearson’s chi-square test (sex, N with hypertension at baseline, ischaemic stroke subtype), and a Fisher’s exact test (N with depression, N with mRS score ≥ 2, stroke side, N with T2DM at baseline, N with ischaemic heart disease at baseline).

| Variable | Non-Fatigued | Fatigued | *p*-Value<br>(2-tailed) | Effect Size |
| --- | --- | --- | --- | --- |
| *N* (*N* female) | 18 (5) | 45 (17) | .564 | .10<sup>a</sup> |
| Age – *Mdn* (IQR) | 67.50 (16) | 71.00 (19) | .254 | .14<sup>b</sup> |
| PHQ-9 – *Mdn* (IQR) | 1 (2) | 4 (6) | <.001 | .52<sup>b</sup> |
| *N* with depression (PHQ-9 ≥ 5) | 0 | 19 | .001 | .42<sup>a</sup> |
| NIHSS at baseline – *Mdn* (IQR) | 2 (3) | 2 (3) | .619 | .06<sup>b</sup> |
| *N* with mRS ≥ 2 | 4 | 15 | .545 | .12<sup>a</sup> |
| Stroke side (*N* left, right, bilateral) | 8, 9, 1 | 17, 27, 1 | .520 | .11<sup>a</sup> |
| Lesion volume (ml) – *Mdn* (IQR) | 1.90 (11.93) | 1.40 (6.83) | .534 | .09<sup>b</sup> |
| GAD-7 – *Mdn* (IQR) | 0 (2) | 3 (3) | .011 | .32<sup>b</sup> |
| *N* with T2DM at baseline | 0 | 12 | .013 | .31<sup>a</sup> |
| *N* with ischaemic heart disease at baseline | 1 | 4 | .999 | .06<sup>a</sup> |
| *N* with hypertension at baseline | 9 | 27 | .576 | .09<sup>a</sup> |
| Education (in years) at baseline – *Mdn* (IQR) | 12.50 (4) | 11.00 (5) | .422 | .10<sup>b</sup> |
| WAIS-III Digit span *z*-score – *M* (*SD*) | -0.13 (0.82) | -0.03 (0.90) | .691 | .11<sup>c</sup> |
| HVLT-R *z*-Score – *M* (*SD*) | 0.28 (1.18) | 0.09 (1.10) | .550 | .17<sup>c</sup> |
| ROCF delay *z*-score – *M* (*SD*) | 0.004 (1.20) | -0.16 (1.22) | .641 | .13<sup>c</sup> |
| Cogstate™ detection *z*-score – *M* (*SD*) | 0.19 (0.31) | -0.15 (0.55) | .017 | .76<sup>c</sup> |
| Cogstate™ identification *z*-score – *Mdn* (IQR) | 0.05 (0.70) | -0.12 (0.60) | .773 | .04<sup>b</sup> |
| Cogstate™ one-back *z*-score – *Mdn* (IQR) | -0.57 (1.45) | -0.57 (1.73) | .874 | .02<sup>b</sup> |

*Note.* All measures were recorded at 3 months post-stroke unless otherwise stated. PHQ-9 = Patient Health Questionnaire<sup>48</sup>; IQR = interquartile range; NIHSS = National Institutes of Health Stroke Scale<sup>52</sup>; mRS = Modified Rankin Scale<sup>53</sup>; GAD-7 = Generalised Anxiety Disorder questionnaire<sup>54</sup>; T2DM = type 2 diabetes mellitus; WAIS-III = Wechsler Adult Intelligence Scale 3<sup>rd</sup> Edition<sup>106</sup>; HVLT-R = Hopkins Verbal Learning Test Revised for memory recall<sup>107</sup>; ROCF delay = Rey-Osterrieth Complex Figure Delay task<sup>108</sup>.
<sup>a</sup> Cramer's *V* statistic. <sup>b</sup> Correlation coefficient (*r*). <sup>c</sup> Cohen's delta (*d*) statistic.

### Imaging data acquisition and pre-processing

A Siemens 3T Skyra scanner (Erlangen, Germany) with a 12-channel head coil was used to conduct neuroimaging. A high-resolution magnetization-prepared rapid acquisition with gradient echo (MP-RAGE) volume was acquired, with 160 sagittal slices and 1 mm isotropic voxels, repetition time (TR) = 1900 ms, echo time (TE) = 2.55 ms, 9° flip angle, 100% field of view in the phase direction and 256 × 256 acquisition matrix. High-resolution 3D SPACE-FLAIR images were also collected, comprising of 160 sagittal 1 mm thick slices, TR = 6000 ms, TE = 380 ms, 120° flip angle, 100% field of view in the phase direction, 256 × 256 acquisition matrix, and 512 × 512 reconstruction matrix. Resting-state data were obtained via axial oriented, interleaved slices with 3 mm isotropic voxels, 3 mm slice gap, TR = 3000 ms, TE = 30 ms and 85° flip angle, 100% field of view in phase direction and 72 × 72 acquisition matrix. During resting-state data acquisition, participants were instructed to keep their eyes closed.

SPM8^55^ was used to pre-process functional images. These images were slice-time corrected with the middle slice serving as a reference. Images were co-registered to the high-resolution structural image and six-parameter rigid body realignment was performed.

Manually traced lesions generated via the high-resolution FLAIR image were visually inspected and verified by a stroke neurologist. To improve tissue segmentation and preserve lesion size,^56, 57^ a binary lesion mask was generated and used for segmentation and normalisation to the MNI152 template as well as with the Clinical Toolbox SPM extension.^58^ These tissue segmentations were manually examined to maintain quality assurance and functional images were smoothed with an 8 mm full width half maximum Gaussian kernel. MRIcron software^59^ was used to visualise lesion overlap and lesion volume was estimated using SPM12.^60^

The REST software,^61^ which has in-built fast Fourier transform functions, was used to compute the fractional amplitude of low-frequency fluctuation (fALFF) values from detrended data. Relative to the entire frequency range (0-0.25 Hz), the ratios of power were calculated in the 0.01 – 0.08 Hz band, the slow-5 band (0.01 – 0.027Hz), and the slow-4 (0.027 – 0.073 Hz) band.

### Data handling

Data screening analyses were conducted as per Tabachnick and Fidell^62^ to determine the amount and type of missing data present and to inspect dependent variables for violations of normality and homogeneity of variance. Participants were divided into two groups based on their scoring of item 4 of the PHQ-9, as it has been shown to correspond to the total score of the Fatigue Questionnaire.^50^ Those who scored > 0 were placed in the fatigued group whereas those who scored 0 were placed in the non-fatigued group. All fALFF values were transformed to *z*-values before analyses, which were performed at the whole-brain level.

### Statistical analysis

The two groups were compared on clinical and cognitive variables (Table 1). Effect sizes used for analysis are shown in Supplementary Table I. The alpha level for these group comparisons was set at *p* <.05. Imaging results were considered significant at the voxel level of *p* <.005 and cluster-corrected for multiple comparisons level of *p*_*FDR*_ <.05. All data screening and group comparison analyses were conducted via IBM SPSS Statistics (Version 26).

### Primary fALFF analyses

A generalised linear model analysis was conducted via SPM12 to compare the fALFF maps for fatigued and non-fatigued participants on a voxel-wise basis. Participant’s age, sex, and NIHSS at baseline were set as covariates. This group analysis was conducted in the combined 0.01 – 0.08 Hz band as well as the slow-5 (0.01 - 0.027 Hz) and slow-4 (0.027 - 0.073 Hz) frequencies.

### Secondary analyses

Post-hoc analyses were conducted to ensure that results from the primary fALFF analyses were not driven by confounding factors. Firstly, to ensure that resting-state results were not due to the lesion overlap with the significant cluster, the main fatigued vs non-fatigued 0.01 - 0.08 Hz analysis was repeated with participants who demonstrated lesion overlap with significant clusters removed.

Secondly, to address the confound of depression, fatigued participants were separated into two groups according to depression status (PHQ-9 score ≥ 5). The fALFF maps of depressed and non-depressed fatigued participants were then compared in the 0.01 – 0.08 Hz band on a voxel-wise basis using a generalised linear model analysis with age, sex, and NIHSS at baseline as covariates to determine whether fatigue and depression have distinct or overlapping neural correlates.

### Data availability

The data that support the findings of this study are available on reasonable request from the corresponding authors. All requests for raw and analysed data will be reviewed by the CANVAS investigators to determine whether the request is subject to any intellectual property or confidentiality obligations.

## Results

### Data screening

Total missing data for all participants across all variables was 1.9%. Little’s Missing Completely at Random (MCAR) test (MCAR, χ^2^(58) = 74.89, *p* = .067), indicated that the missingness could be disregarded. Inspection of all dependent variables revealed several violations of normality and homogeneity of variance (Supplementary Table II and III). For these variables, a non-parametric Mann-Whitney *U* test was employed for group comparisons.

### Behavioural results

Data from 63 stroke patients (22 women) aged 30 to 89 years (*M* = 67.51, *SD* = 13.39) from the CANVAS study were analysed. Each participant had behavioural data available at 3 months post-stroke and a full set of imaging data. Forty-five participants (71.4%) reported experiencing PSF (PHQ-9 item 4 > 0) and were assigned to the fatigued group while the rest were classified as non-fatigued. Behavioural results are presented in Table 1 (see Supplementary Tables IV-VI for full inferential statistics). Notably, fatigue status was significantly associated with PHQ-9 score, depression status, and GAD-7 score, which exhibited a large, medium-to-large, and medium effect size, respectively^63^. Given that GAD-7 and PHQ-9 are highly correlated,^54, 64, 65^ we focused on the depression group analysis. T2DM diagnosis and Cogstate™^66^ detection task scores were also significantly associated with fatigue status, where the former constituted a medium effect, and the latter a medium-to-large effect size.^63^

### Analysis of ischaemic stroke subtype

No participants were classified as having experienced a total anterior cerebral infarction (TACI). While there appeared to be some quantitative differences in the proportion of fatigued and non-fatigued participants with posterior cerebral infarctions (POCI), partial anterior cerebral infarctions (PACI), and lacunar cerebral infarctions (LACI; Fig. 1), a Fisher’s exact test found no significant differences between groups, *p* = .686, two-tailed, *V* = .11.

**Figure 1.**
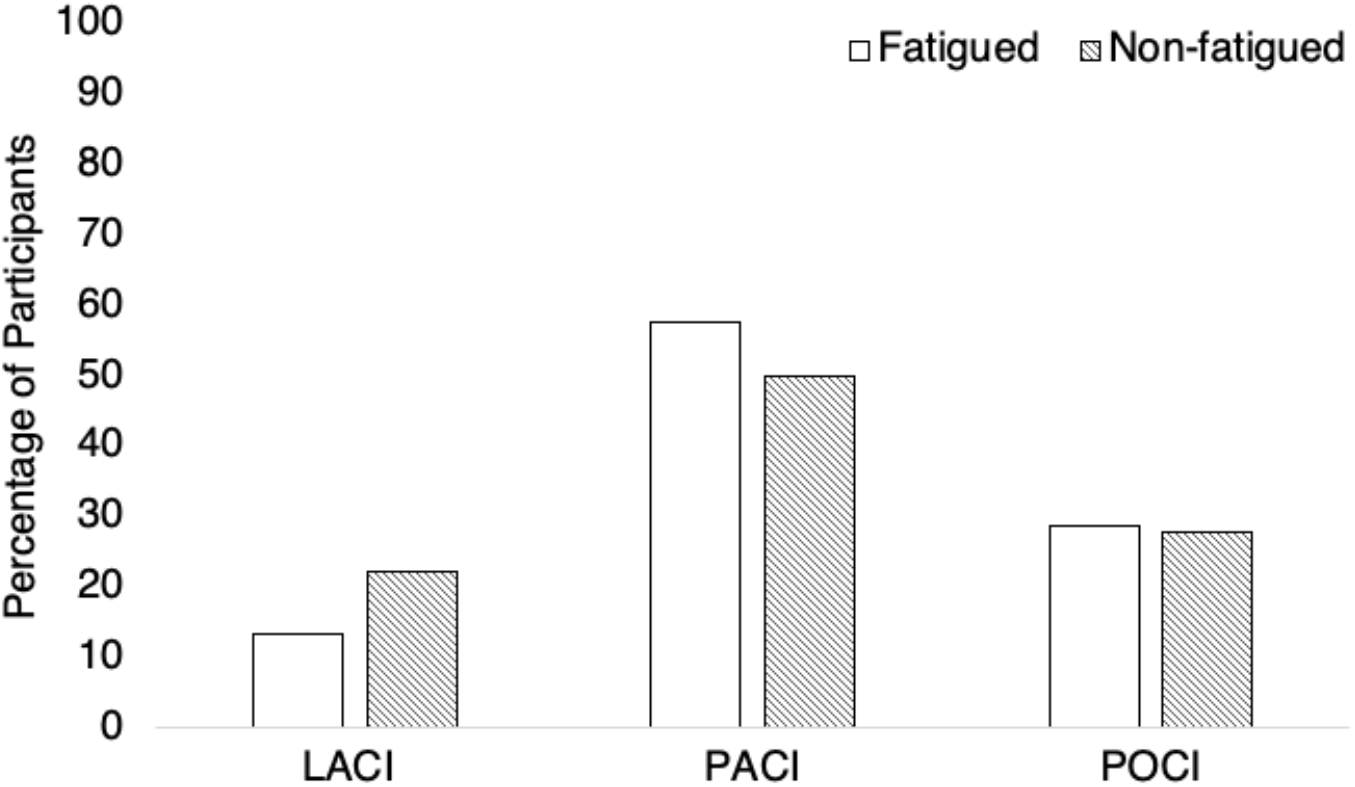
Distribution of Ischaemic Stroke Subtypes Across Fatigued vs Non-Fatigued Groups. *Note*. LACI = lacunar cerebral infarction; PACI = partial anterior cerebral infarction; POCI = posterior cerebral infarction.

### Analysis of lesion location

For all participants, lesions were widely distributed with more right-hemisphere lesions (Fig. 2A). Within non-fatigued participants there was no significant lesion overlap (Fig. 2B). Fatigued participants demonstrated some overlap in the right middle cerebral artery, as well as surrounding areas in the right hemisphere such as the thalamus and striatocapsular regions (Fig. 2C). However, given that the magnitude of overlap was at most 6 participants, i.e. only 13.3% of the fatigued group, there does not appear to be a strong association with fatigue.

**Figure 2.**
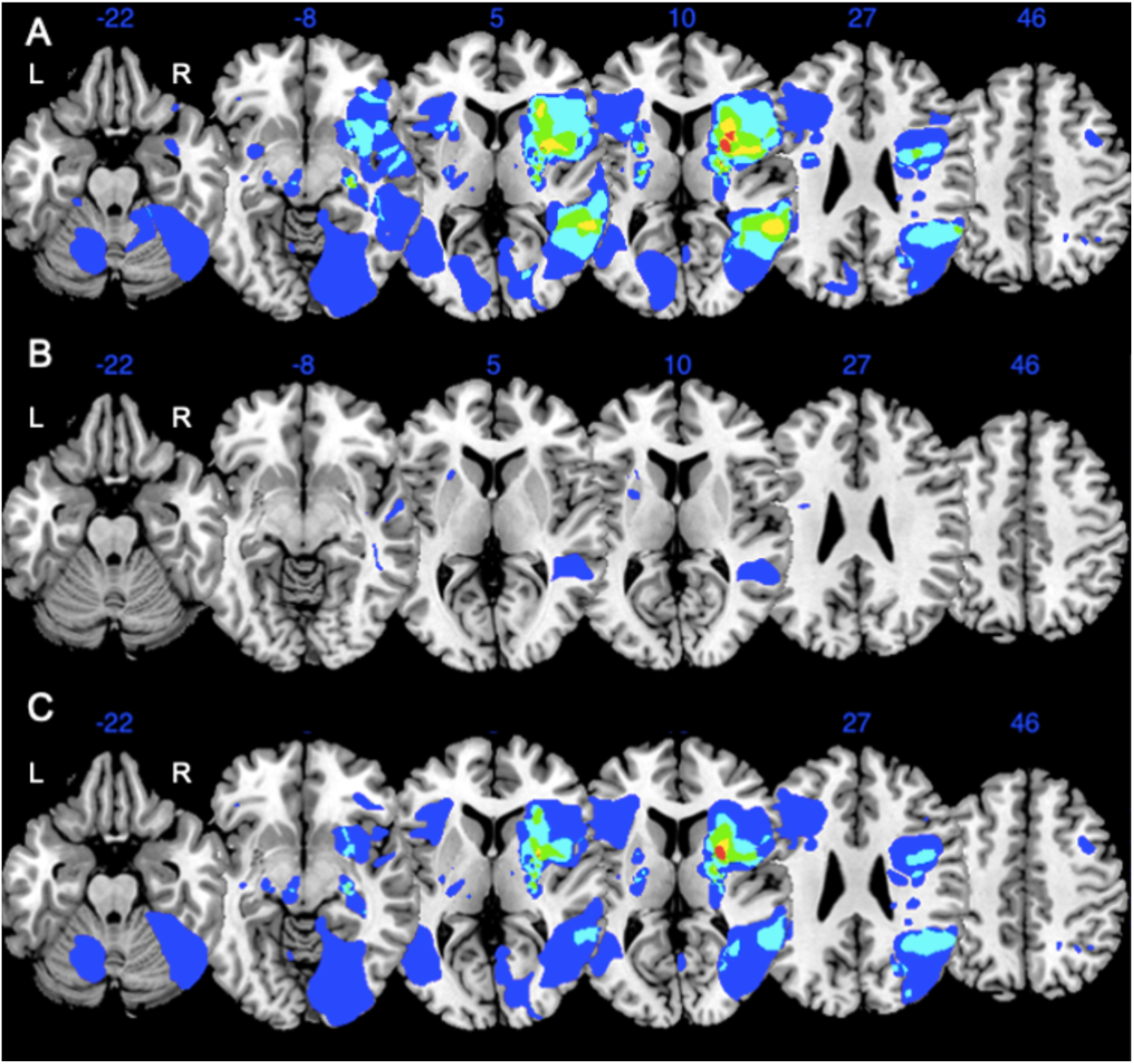
Comparison of Lesion Overlap in Non-Fatigued and Fatigued Groups. A. Lesion Overlap Map for All Participants (N = 63). B. Non-fatigued participants (N = 18). C. Fatigued participants (N = 45). The colours reflect the number of participants with overlapping lesions (blue = 2 participants, light blue = 3 participants, green = 4 participants, yellow = 5 participants, red = 6 participants).

### fALFF in 0.01–0.08 Hz band

Significant group differences were found in both frontal and posterior regions. Namely, fatigued participants demonstrated significantly higher activity in the bilateral medial prefrontal cortex (mPFC) and significantly lower activity in the bilateral calcarine cortex and left lingual gyrus than non-fatigued participants (Table 2; Fig. 3).

**Table 2.**
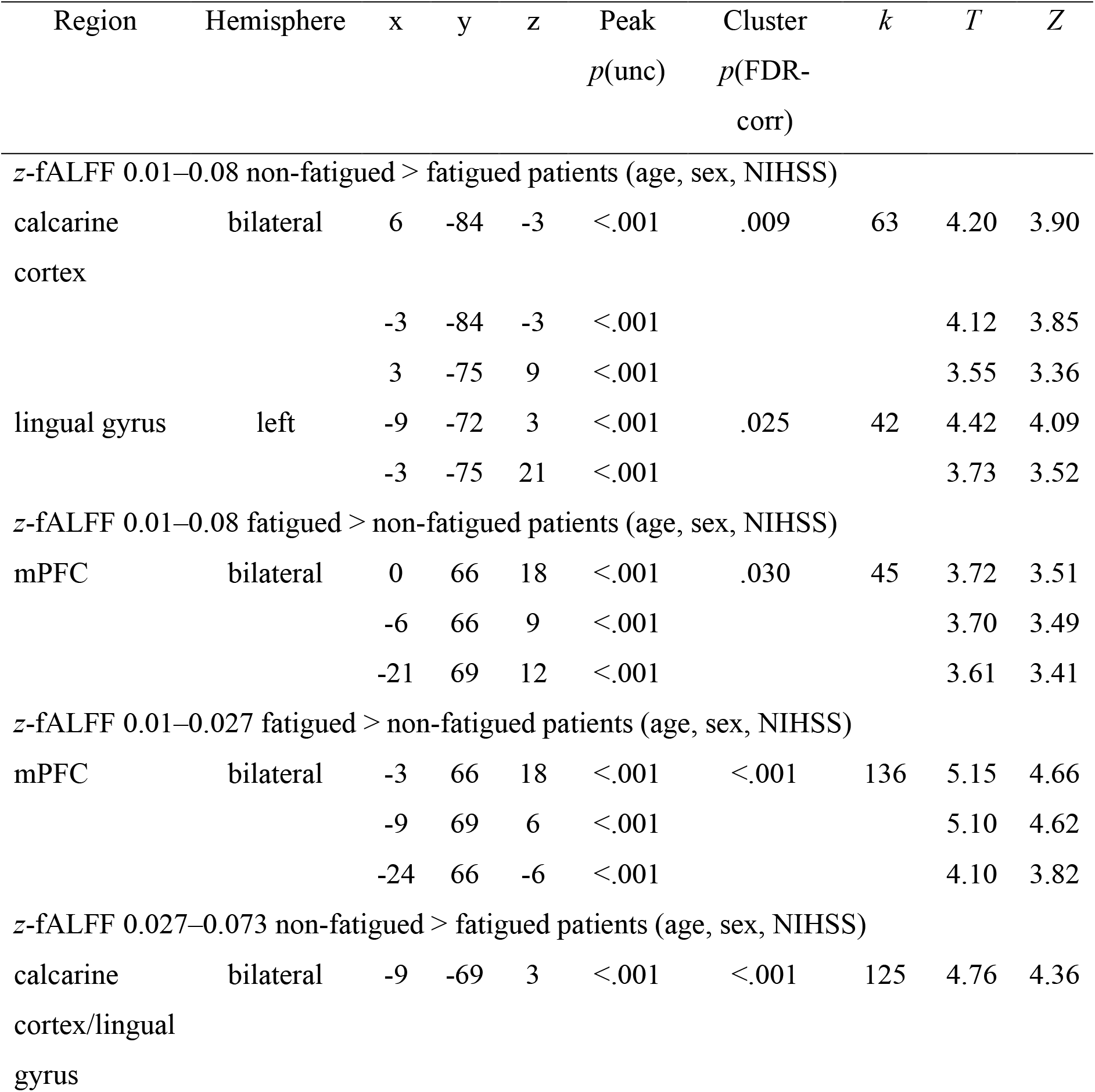
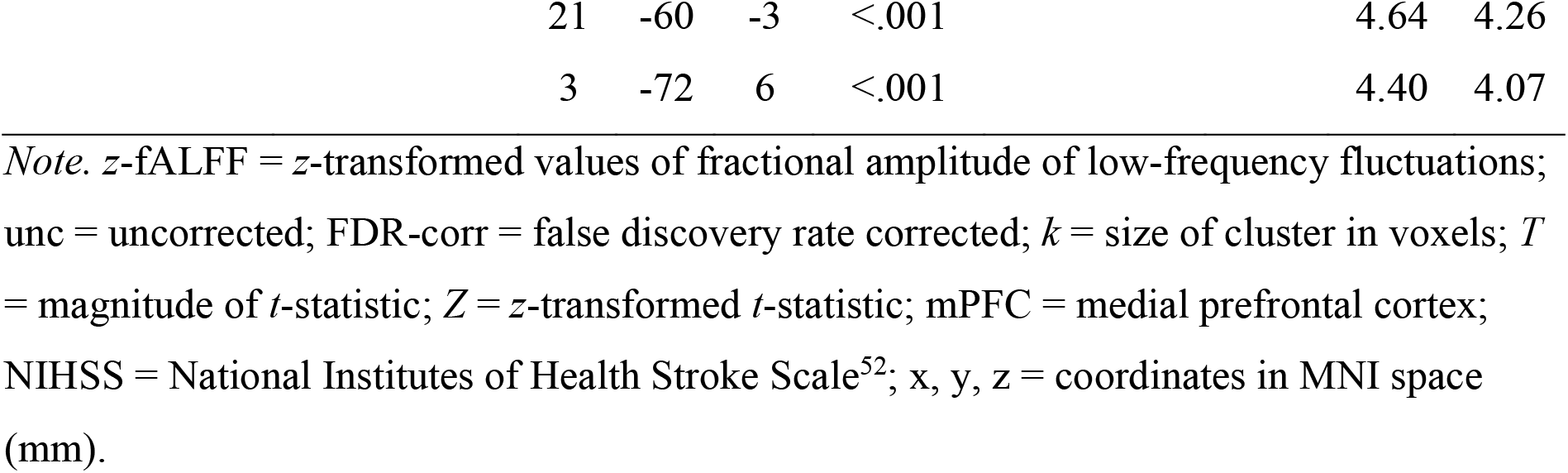
Significant z-fALFF Results for Fatigued vs Non-Fatigued in the 0.01-0.08 Hz and Slow-4 and Slow-5 Bands

**Figure 3.**
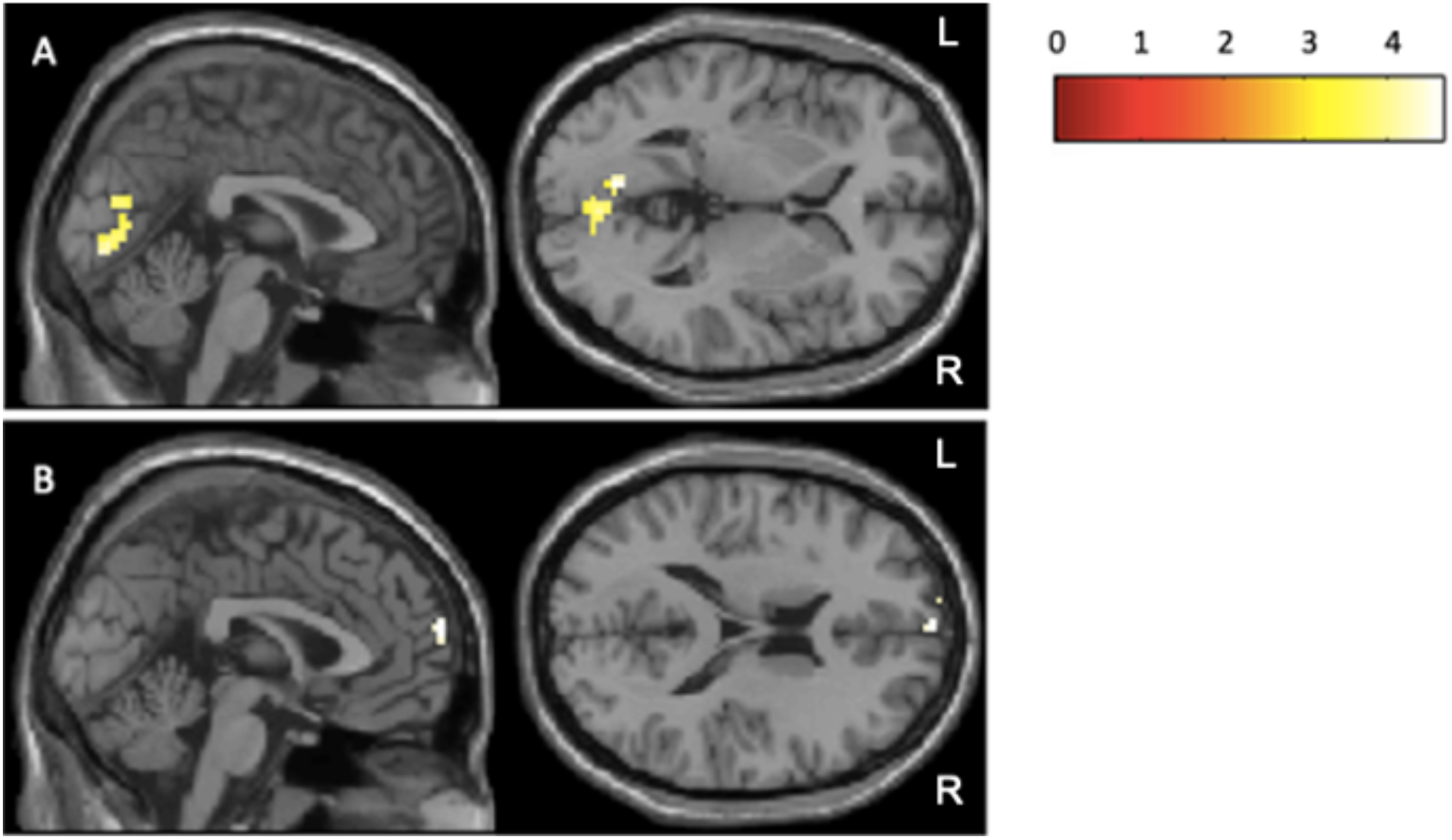
Main Results in fALFF for Fatigued vs Non-Fatigued in the 0.01 – 0.08 Hz Band. A. Results for the non-fatigued > fatigued contrast. B. Results for the fatigued > non-fatigued contrast. The colour bar represents the magnitude of the t-statistic computed from the analysis.

### fALFF in slow-5 (0.01–0.027 Hz) and slow-4 (0.027–0.073 Hz) bands

A significant group difference was found in the slow-5 band in the mPFC where fatigued participants demonstrated significantly higher activity than non-fatigued participants (Table 2). In the slow-4 band, a group difference was identified in the bilateral calcarine cortex/lingual gyrus, with fatigued participants showing significantly lower activity than non-fatigued participants. A cluster (*k* = 27) in the slow-4 band in the left superior parietal cortex was significant at the voxel level (*p* = .010, *T* = 4.88, *Z* = 4.45, *MNI*: −21, −51, 66), but not at the cluster-corrected level, *p*_*FDR*_ = .139. Opposite contrasts for both the slow-5 and slow-4 bands showed no suprathreshold clusters.

### Secondary analyses

#### Analysis of lesion location with respect to significant clusters

Only one participant had a lesion overlapping with the significant calcarine cluster in the fatigued vs non-fatigued analysis in the 0.01 – 0.08 Hz band (Supplementary Fig. I). When this participant was excluded from analysis, the same clusters in the mPFC and calcarine cortex/lingual gyrus were identified. The calcarine cluster specifically remained significant at both voxel- and cluster-corrected levels (Supplementary Table VII).

#### Depressed fatigued vs non-depressed fatigued analysis in 0.01–0.08 Hz band

Firstly, while all non-fatigued participants were not depressed, only 42% (n = 19) of fatigued participants exhibited concomitant depression. A significant group difference was found in the left dorsolateral prefrontal cortex (dlPFC; Supplementary Table VIII; Supplementary Fig. II), where the depressed participants exhibited significantly higher activity compared to those who were not depressed. In the opposite contrast, two clusters were significant at the voxel level, one in the left cerebral white matter (*k* = 22, *p* = .025, *T* = 3.91, *Z* = 3.58, *MNI*: −18, 33, 6) and the other in the right dlPFC (*k* = 20, *p* = .032, *T* = 3.66, *Z* = 3.37, *MNI*: 21, 42, 21), yet neither were significant after cluster correction: *p*_*FDR*_ = .394. The overlay of the significant clusters from this secondary analysis and the main fatigued vs non-fatigued analysis is shown in Supplementary Fig. III and demonstrates no overlap between the clusters associated with fatigue (in the main fatigued vs non-fatigued analysis) and depression within the fatigued group (in the depressed fatigued vs non-depressed fatigued analysis).

## Discussion

We aimed to identify neural correlates of PSF via a whole brain connectivity approach. No differences in stroke characteristics (stroke type, lesion location and volume) were observed between fatigued and non-fatigued stroke participants. We found fatigued participants demonstrated significantly increased neural activity in the mPFC compared to non-fatigued, mostly attributable to slow-5 oscillations, demonstrating dysfunction of the *a priori* hypothesised FST network. Fatigued participants additionally showed significant hypoactivity in the calcarine cortex and lingual gyrus driven by slow-4 oscillations. This latter association has not been previously described in the PSF literature but is consistent with previous neuroimaging studies of fatigue in non-stroke populations.^67-70^

### Resting-state correlates

#### Hyperactivity in the medial prefrontal cortex

Concordant with the hypothesis predicting a significant difference in resting-state activity between those with and without PSF, fatigued participants demonstrated significantly higher fALFF activity in the mPFC in comparison to non-fatigued participants. This hyperactivity was mostly linked to slow-5 oscillations, which aligns well with reports that slow-5 fALFF are more sensitive to signals from the cortical regions.^71^ Although the mPFC specifically has not been strongly linked to PSF, these findings are consistent with observations of fatigue in other clinical populations. Namely, our results are consistent with findings of perturbed PFC activity, abnormal mPFC glucose metabolism, and altered functional connectivity between the basal ganglia and mPFC in fatigued MS patients^72, 73^ and altered FST activity and functional connectivity in fatigued Parkinson’s, TBI, and CFS patients.^40-43, 74^ Furthermore, the observed mPFC dysfunction substantiates findings that cognitive fatigue is linked to cortico-striatal network dysfunction, particularly between the basal ganglia and PFC, in both clinical and general populations.^75^

While we did not specifically observe aberrant resting-state activity in the thalamus or striatum of fatigued participants, the mPFC is a major cortical node in FST circuitry,^34, 76^ and thus our findings of frontal dysfunction in fatigued participants provide evidence for the role of the FST network in PSF. Given the FST network’s multifaceted role in mediating numerous cognitive, motor, and affective processes,^77, 78^ the observed hyperactivation in the mPFC could be a reflection of the increased cerebral effort required by fatigued participants in comparison to their non-fatigued counterparts, to recruit and facilitate these processes, even in the absence of a task. This increased cerebral effort may then be manifested as the subjective feeling of fatigue; a notion that aligns well with sleep restriction literature that reports increased prefrontal and parietal activity observed in response to cognitive demands following total sleep deprivation.^79^ However, further study is required to consolidate and extend upon the relationship between PSF and mPFC hyperactivity.

### Hypoactivity in the calcarine cortex and lingual gyrus

The left lingual gyrus and bilateral calcarine cortex exhibited significantly lower fALFF activity in those with PSF compared to non-fatigued participants. This difference was largely attributed to slow-4 oscillations, corroborating the concept that slow-4 fALFF may reflect signals from subcortical regions.^71^ In contrast to our findings of abnormal mPFC activity which aligned well with FST dysfunction specified in previous literature, these particular loci have not been previously linked with PSF and thus constitute novel contributions to the PSF literature.

Given the proximity of both the calcarine cortex and lingual gyrus to the primary visual cortex and their roles in encoding visual input such as complex images,^80^ landmarks,^81^ words,^82^ and facial expressions,^83^ the irregular activity observed in fatigued participants could suggest a relationship between PSF and abnormalities in visual functioning. For example, there exists a body of research in non-stroke populations linking age-related dysfunction in the calcarine cortex and lingual gyrus with impairment of the well-described alerting function of light during daytime.^84, 85^ Thus, given the observed occipital dysfunction, it is plausible that this alerting mechanism could be disrupted in PSF, thus resulting in decreased alertness, increased fatigue, and a disrupted circadian rhythm.^85^

Additionally, taken together with our findings of mPFC dysfunction, aberrant resting-state activity in the calcarine cortex and lingual gyrus could indicate frontal-occipital network dysfunction in PSF’s pathophysiology. The frontal-occipital network, connected via a large association tract, the inferior fronto-occipital fasciculus,^86^ facilitates the integration of visual information throughout the cortex and has been implicated in higher-order processes such as visual selective attention,^87^ lexical and semantic processing, and visual memory.^88^ Therefore, our observation of abnormal resting-state activity in both frontal and occipital regions could contribute to disruptions in processing: a notion that is consistent with the fact that approximately 65% of stroke survivors experience some form of post-stroke visual impairment such as visual inattention, visual field loss, or ocular motility problems.^89^

Indeed, given the high functional integration between prefrontal and occipital cortices,^86^ the hyperactivity in the mPFC may be serving some compensatory function for decreased occipital lobe activity, whereby cerebral effort in the mPFC increases to account for underperforming primary visual areas, which then may manifest as the subjective feeling of fatigue. Appelros and colleagues reported an association between PSF and visual impairment,^22^ and several authors have outlined a bidirectional relationship between pathological fatigue and vision,^90-93^ where those with visual impairment exhibit more severe fatigue than persons with normal sight and those with CFS often exhibit concomitant visual deficits. As such, further investigation is required to clarify the relationship between these two post-stroke sequelae and determine whether these same resting-state occipital clusters are linked to PSF in the absence of visual impairments.

### PSF and stroke characteristics

Despite previous literature linking PSF with stroke subtype^24-26, 28, 31, 32^ and lesion location^18-20, 29^ neither were found to be associated with fatigue in the current study. This is consistent with prior reports finding that PSF does not depend on stroke location.^21-23^ The null association between fatigue status, lesion volume, and stroke side appears consistent with prior PSF literature.^15, 67, 94^ Importantly, while an association between PSF and the posterior circulation territory was not found via lesion or ischaemic stroke subtype analysis, both the calcarine cortex and lingual gyrus are supplied by the posterior cerebral artery,^95^ and thus our findings of abnormal resting-state activity in these regions could, by inference, add weight to the hypothesised link between PSF and the posterior circulation system. However, replication and further investigation are required to clarify this relationship and extend upon these findings.

### Association with depression and anxiety symptoms

Whilst PHQ-9 and GAD-7 scores were significantly associated with fatigue status, we demonstrated dissociable effects of PSF and depression. Firstly, >50% of participants who presented with PSF did not have depression, reinforcing the notion that the two sequelae can exist independently.^19^ Secondly, after observing the resting-state neural correlates of depression within fatigued participants in secondary analyses, depression was linked to increased fALFF in the left dlPFC. The dlPFC has been previously implicated in depression and post-stroke depression in particular. Namely, depression-related lesions in the left dlPFC^96^, increases in dlPFC grey matter volume,^97^ altered resting-state functional connectivity of the dlPFC,^98-101^ and aberrant ALFF and fALFF activity in the left dlPFC.^102-104^

Given the significant clusters reflecting depression and PSF demonstrated no overlap, the brain regions attributable to fatigue and depression post-stroke were effectively differentiated.

### Limitations and future directions

The most prominent limitation of this study is that we measured PSF via a single item from a subjective depression severity questionnaire. As fatigue is one of the nine key diagnostic symptoms for major depressive disorder,^49^ the two post-stroke complications are highly comorbid and difficult to disentangle. Therefore, while secondary analyses successfully distinguished the resting-state neural correlates of the two sequelae, we cannot conclusively rule out the influence that depression may exert on our results nor the possibility that some form of depression-fatigue interaction may be present. Above all, this limitation highlights the growing need for a stroke-validated measure and definition of PSF in the literature, which should be addressed in future research.

This study also did not attempt to distinguish between different types of fatigue. Though the observation of significantly lower Cogstate™ detection task scores in fatigued participants could plausibly implicate the observed clusters in mental fatigue manifested as delayed processing speed, one cannot discount the possible contribution of physical fatigue to performance, especially given the task’s psychophysical nature. Future studies should adopt a more multifaceted approach to fatigue.

While significant neural correlates were identified, we cannot conclude causality; that is, whether PSF is a result of aberrant activity in the mPFC, calcarine cortex, and lingual gyrus, or whether PSF itself causes dysfunction in these brain regions.

Furthermore, as we examined PSF at 3 months post-stroke, our findings do not provide insight into PSF’s progression over time, particularly regarding the stability of its resting-state neural correlates. Given the distinction between early and late fatigue regarding their definitions and associated factors,^105^ PSF could plausibly reflect differing resting-state neural correlates at different time points. This limitation should be addressed in future research via longitudinal studies that assess the nature and stability of PSF’s resting-state neural correlates over time.

Lastly, our sample was primarily composed of individuals who had experienced minor or moderate strokes. Future research should investigate if these findings are generalisable to patients with more severe strokes.

## Conclusion

We found that hyperactivity in the prefrontal and hypoactivity in the occipital cortices is associated with PSF, beyond the influence of confounding demographic or neurological factors. In turn, these findings may implicate dysfunction in several large-scale brain networks such as the FST and frontal-occipital network, in the pathophysiology of PSF. We found no association between PSF, lesion location and subtype of ischaemic stroke. Our novel findings unearth several avenues for future PSF research, where the identified loci and their relationship with PSF can be further explored with seed-based neuroimaging and serve as targets for therapeutic brain stimulation.

## References

1. Staub F, Bogousslavsky J. Fatigue after stroke: a major but neglected issue. Cerebrovasc Dis. 2001;12(2):75–81.

2. Röding J, Lindström B, Malm J, Ohman A. Frustrated and invisible--younger stroke patients’ experiences of the rehabilitation process. Disabil Rehabil. 2003;25(15):867–874.

3. Naess H, Nyland H. Poststroke fatigue and depression are related to mortality in young adults: a cohort study. BMJ Open. 2013;3(3): e002404.

4. Christensen D, Johnsen SP, Watt T, Harder I, Kirkevold M, Andersen G. Dimensions of post-stroke fatigue: a two-year follow-up study. Cerebrovasc Dis. 2008;26(2):134–141.

5. Lerdal A, Bakken LN, Kouwenhoven SE, et al. Poststroke fatigue--a review. J Pain Symptom Manage. 2009;38(6):928–949.

6. Nadarajah M, Goh HT. Post-stroke fatigue: a review on prevalence, correlates, measurement, and management. Top Stroke Rehabil. 2015;22(3):208–220.

7. Aarnes R, Stubberud J, Lerdal A. A literature review of factors associated with fatigue after stroke and a proposal for a framework for clinical utility. Neuropsychol Rehabil. 2020:30:1449–1476.

8. Lerdal A, Wahl A, Rustøen T, Hanestad BR, Moum T. Fatigue in the general population: a translation and test of the psychometric properties of the Norwegian version of the fatigue severity scale. Scand J Public Health. 2005;33(2):123–130.

9. Cantor JB, Ashman T, Gordon W, et al. Fatigue after traumatic brain injury and its impact on participation and quality of life. J Head Trauma Rehabil. 2008;23(1):41–51.

10. Lamers F, Hickie I, Merikangas KR. Prevalence and correlates of prolonged fatigue in a U.S. sample of adolescents. Am J Psychiatry. 2013;170(5):502–510.

11. Galland-Decker C, Marques-Vidal P, Vollenweider P. Prevalence and factors associated with fatigue in the Lausanne middle-aged population: a population-based, cross-sectional survey. BMJ Open. 2019;9(8):e027070.

12. Kutlubaev MA, Duncan FH, Mead GE. Biological correlates of post-stroke fatigue: a systematic review. Acta Neurol Scand. 2012;125(4):219–227.

13. Hinkle JL, Becker KJ, Kim JS, et al. Poststroke fatigue: emerging evidence and approaches to management: a scientific statement for healthcare professionals from the american heart association. Stroke. 2017;48(7):e159–e170.

14. Kennedy C, Kidd L. Interventions for post-stroke fatigue: a Cochrane review summary. Int J Nurs Stud. 2018;85:136–137.

15. Ponchel A, Bombois S, Bordet R, Hénon H. Factors associated with poststroke fatigue: a systematic review. Stroke Res Treat. 2015:347920.

16. Bamford J, Sandercock P, Dennis M, Burn J, Warlow C. Classification and natural history of clinically identifiable subtypes of cerebral infarction. Lancet. 1991;337(8756):1521–1526.

17. Markus HS, van der Worp HB, Rothwell PM. Posterior circulation ischaemic stroke and transient ischaemic attack: diagnosis, investigation, and secondary prevention. Lancet Neurol. 2013;12(10):989–998.

18. Pittock SJ, Meldrum D, Hardiman O, Thornton J, Brennan P, Moroney JT. The Oxfordshire Community Stroke Project classification: correlation with imaging, associated complications, and prediction of outcome in acute ischemic stroke. J Stroke Cerebrovasc Dis. 2003;12(1):1–7.

19. Naess H, Nyland HI, Thomassen L, Aarseth J, Myhr KM. Fatigue at long-term follow-up in young adults with cerebral infarction. Cerebrovasc Dis. 2005;20(4):245–50.

20. Chen K, Marsh EB. Chronic post-stroke fatigue: It may no longer be about the stroke itself. Clin Neurol Neurosurg. 2018;174:192–197.

21. Choi-Kwon S, Han SW, Kwon SU, Kim JS. Poststroke fatigue: characteristics and related factors. Cerebrovasc Dis. 2005;19(2):84–90.

22. Appelros P. Prevalence and predictors of pain and fatigue after stroke: a population-based study. Int J Rehabil Res. 2006;29(4):329–333.

23. Schepers VP, Visser-Meily AM, Ketelaar M, Lindeman E. Poststroke fatigue: course and its relation to personal and stroke-related factors. Arch Phys Med Rehabil. 2006;87(2):184–188.

24. Mutai H, Furukawa T, Houri A, Suzuki A, Hanihara T. Factors associated with multidimensional aspect of post-stroke fatigue in acute stroke period. Asian J Psychiatr. 2017;26:1–5.

25. Snaphaan L, van der Werf S, de Leeuw FE. Time course and risk factors of post-stroke fatigue: a prospective cohort study. Eur J Neurol. 2011;18(4):611–617.

26. Tang WK, Liang HJ, Chen YK, et al. Poststroke fatigue is associated with caudate infarcts. J Neurol Sci. 2013;324(1-2):131–135.

27. Manes F, Paradiso S, Robinson RG. Neuropsychiatric effects of insular stroke. J Nerv Ment Dis. 1999;187(12):707–712.

28. Naess H, Lunde L, Brogger J. The effects of fatigue, pain, and depression on quality of life in ischemic stroke patients: the Bergen Stroke Study. Vasc Health Risk Manag. 2012;8:407–413.

29. Kutlubaev MA, Shenkin SD, Farrall AJ, et al. CT and clinical predictors of fatigue at one month after stroke. Cerebrovasc Dis Extra. 2013;3(1):26–34.

30. Nouh A, Remke J, Ruland S. Ischemic posterior circulation stroke: a review of anatomy, clinical presentations, diagnosis, and current management. Front Neurol. 2014;5:30.

31. Tang WK, Chen YK, Mok V, et al. Acute basal ganglia infarcts in poststroke fatigue: an MRI study. J Neurol. 2010;257(2):178–182.

32. Wei C, Zhang F, Chen L, Ma X, Zhang N, Hao J. Factors associated with post-stroke depression and fatigue: lesion location and coping styles. J Neurol. 2016;263:269-76. Retraction in: Wei C, Zhang F, Chen L, Ma X, Zhang N, Hao J. J Neurol. 2018;265:451.

33. Turner IF, Ward NS, Kuppuswamy A. Fatigue, effort perception and central activation failure in chronic stroke survivors: a TMS and fMRI investigation. bioRxiv. 2019:821959.

34. Alexander GE, DeLong MR, Strick PL. Parallel organization of functionally segregated circuits linking basal ganglia and cortex. Annu Rev Neurosci. 1986;9(1):357–381.

35. Visser MM, Maréchal B, Goodin P, et al. Predicting modafinil-treatment response in poststroke fatigue using brain morphometry and functional connectivity. Stroke. 2019;50(3):602–609.

36. Zaini WH, Giuliani F, Beaulieu C, Kalra S, Hanstock C. Fatigue in multiple sclerosis: assessing pontine involvement using proton MR spectroscopic imaging. PLoS One. 2016;11(2):e0149622.

37. Basu N, Murray AD, Jones GT, Reid DM, Macfarlane GJ, Waiter GD. Neural correlates of fatigue in granulomatosis with polyangiitis: a functional magnetic resonance imaging study. Rheumatology (Oxford). 2014;53(11):2080–2087.

38. Berginström N, Nordström P, Ekman U, et al. Using functional magnetic resonance imaging to detect chronic fatigue in patients with previous traumatic brain injury: changes linked to altered striato-thalamic-cortical functioning. J Head Trauma Rehabil. 2018;33(4):266–274.

39. Kohl AD, Wylie GR, Genova HM, Hillary FG, Deluca J. The neural correlates of cognitive fatigue in traumatic brain injury using functional MRI. Brain Inj. 2009;23(5):420–432.

40. Kwak Y, Peltier S, Bohnen N, Müller M, Dayalu P, Seidler R. Altered resting state cortico-striatal connectivity in mild to moderate stage Parkinson’s disease. Original research. Front Syst Neurosci. 2010;4:143.

41. Schifitto G, Deng L, Yeh TM, et al. Clinical, laboratory, and neuroimaging characteristics of fatigue in HIV-infected individuals. J Neurovirol. 2011;17(1):17–25.

42. Wu Q, Inman RD, Davis KD. Fatigue in ankylosing spondylitis is associated with the brain networks of sensory salience and attention. Arthritis Rheumatol. 2014;66(2):295–303.

43. de Lange FP, Kalkman JS, Bleijenberg G, et al. Neural correlates of the chronic fatigue syndrome—an fMRI study. Brain. 2004;127(9):1948–1957.

44. Zou QH, Zhu CZ, Yang Y, et al. An improved approach to detection of amplitude of low-frequency fluctuation (ALFF) for resting-state fMRI: fractional ALFF. J Neurosci Methods. 2008;172(1):137–141.

45. Zuo X-N, Di Martino A, Kelly C, et al. The oscillating brain: complex and reliable. NeuroImage. 2010;49(2):1432–1445.

46. Buzsáki G, Draguhn A. Neuronal oscillations in cortical networks. Science. 2004;304(5679):1926–1929.

47. Brodtmann A, Werden E, Pardoe H, et al. Charting cognitive and volumetric trajectories after stroke: protocol for the Cognition And Neocortical Volume After Stroke (CANVAS) study. Int J Stroke. 2014;9(6):824–828.

48. Kroenke K, Spitzer RL, Williams JB. The PHQ-9: validity of a brief depression severity measure. J Gen Intern Med. 2001;16(9):606–613.

49. American Psychiatric Association. Depressive disorders. In: Diagnostic and statistical manual of mental disorders. American Psychiatric Association; 2013:155–188.

50. Dahl AA, Grotmol KS, Hjermstad MJ, Kiserud CE, Loge JH. Norwegian reference data on the Fatigue Questionnaire and the Patient Health Questionnaire-9 and their interrelationship. Ann Gen Psychiatry. 2020;19(1):60.

51. Goldstein LB, Jones MR, Matchar DB, et al. Improving the reliability of stroke subgroup classification using the Trial of ORG 10172 in Acute Stroke Treatment (TOAST) criteria. Stroke. 2001;32(5):1091–1097.

52. Brott T, Adams HP, Jr., Olinger CP, et al. Measurements of acute cerebral infarction: a clinical examination scale. Stroke. 1989;20(7):864–870.

53. Rankin J. Cerebral vascular accidents in patients over the age of 60. II. Prognosis. Scott Med J. 1957;2(5):200–215.

54. Spitzer RL, Kroenke K, Williams JB, Löwe B. A brief measure for assessing generalized anxiety disorder: the GAD-7. Arch Intern Med. 2006;166(10):1092–1097.

55. The Wellcome Centre for Human Neuroimaging. Statistical Parametric Mapping. Version 8 [software]. 2009 Apr 2 [cited 2020 Oct 30]. Available from https://www.fil.ion.ucl.ac.uk/spm/software/download/

56. Andersen SM, Rapcsak SZ, Beeson PM. Cost function masking during normalization of brains with focal lesions: Still a necessity? NeuroImage. 2010;53(1):78–84.

57. Ripollés P, Marco-Pallarés J, de Diego-Balaguer R, et al. Analysis of automated methods for spatial normalization of lesioned brains. Neuroimage. 2012;60(2):1296–1306.

58. Rorden C, Bonilha L, Fridriksson J, Bender B, Karnath HO. Age-specific CT and MRI templates for spatial normalization. Neuroimage. 2012;61(4):957–965.

59. Rorden C, Karnath H-O, Bonilha L. Improving lesion-symptom mapping. J Cogn Neurosci. 2007;19(7):1081–1088.

60. The Wellcome Centre for Human Neuroimaging. Statistical Parametric Mapping. Version 12 [software]. 2014 Oct 1 [cited 2020 Oct 30]. Available from: https://www.fil.ion.ucl.ac.uk/spm/software/download/

61. Song XW, Dong ZY, Long XY, et al. REST: a toolkit for resting-state functional magnetic resonance imaging data processing. PLoS One. 2011;6(9):e25031.

62. Tabachnick BG, Fidell LS. Using multivariate statistics. Pearson; 2013.

63. Cohen J. Statistical power for the behavioural sciences. Lawrence Erlbaum Associates; 1988.

64. Teymoori A, Gorbunova A, Haghish FE, et al. Factorial structure and validity of depression (PHQ-9) and anxiety (GAD-7) scales after traumatic brain injury. J Clin Med. 2020;9:873.

65. Maroufizadeh S, Omani-Samani R, Almasi-Hashiani A, Amini P, Sepidarkish M. The reliability and validity of the Patient Health Questionnaire-9 (PHQ-9) and PHQ-2 in patients with infertility. Reprod Health. 2019;16(1):137.

66. Cogstate [Internet]. Melbourne: Cogstate; c2020 [cited 2020 Oct 30]. Computerised Cognitive Assessment. Available from https://www.cogstate.com/clinical-trials/computerized-cognitive-assessment/

67. Kluger BM, Zhao Q, Tanner JJ, et al. Structural brain correlates of fatigue in older adults with and without Parkinson’s disease. NeuroImage Clin. 2019;22:101730.

68. Mueller C, Lin JC, Thannickal HH, et al. No evidence of abnormal metabolic or inflammatory activity in the brains of patients with rheumatoid arthritis: results from a preliminary study using whole-brain magnetic resonance spectroscopic imaging (MRSI). Clin Rheumatol. 2020;39(6):1765–1774.

69. Shan ZY, Finegan K, Bhuta S, et al. Brain function characteristics of chronic fatigue syndrome: a task fMRI study. NeuroImage Clin. 2018;19:279–286.

70. Shungu DC, Weiduschat N, Murrough JW, et al. Increased ventricular lactate in chronic fatigue syndrome. III. Relationships to cortical glutathione and clinical symptoms implicate oxidative stress in disorder pathophysiology. NMR Biomed. 2012;25(9):1073–1087.

71. Wang L, Kong Q, Li K, et al. Frequency-dependent changes in amplitude of low-frequency oscillations in depression: A resting-state fMRI study. Neurosci Lett. 2016;614:105–111.

72. Deluca J, Genova H, Hillary F, Wylie G. Neural correlates of cognitive fatigue in multiple sclerosis using functional MRI. J Neurol Sci. 2008;270:28–39.

73. Finke C, Schlichting J, Papazoglou S, et al. Altered basal ganglia functional connectivity in multiple sclerosis patients with fatigue. Mult Scler. 2015;21(7):925–934.

74. Nordin LE, Möller MC, Julin P, Bartfai A, Hashim F, Li T-Q. Post mTBI fatigue is associated with abnormal brain functional connectivity. Sci Rep. 2016;6:21183.

75. Dobryakova E, DeLuca J, Genova HM, Wylie GR. Neural correlates of cognitive fatigue: cortico-striatal circuitry and effort–reward imbalance. J Int Neuropsychol Soc. 2013;19(8):849–853.

76. Dandash O, Pantelis C, Fornito A. Dopamine, fronto-striato-thalamic circuits and risk for psychosis. Schizophr Res. 2017;180:48–57.

77. Tekin S, Cummings JL. Frontal–subcortical neuronal circuits and clinical neuropsychiatry: an update. J Psychosom Res. 2002;53(2):647–654.

78. Avery MC, Krichmar JL. Neuromodulatory systems and their interactions: a review of models, theories, and experiments. Front Neural Circuits. 2017;11:108.

79. Drummond SP, Gillin JC, Brown GG. Increased cerebral response during a divided attention task following sleep deprivation. J Sleep Res. 2001;10(2):85–92.

80. Machielsen WCM, Rombouts SARB, Barkhof F, Scheltens P, Witter MP. fMRI of visual encoding: reproducibility of activation. Hum Brain Mapp. 2000;9(3):156–164.

81. Aguirre GK, D’Esposito M. Topographical disorientation: a synthesis and taxonomy. Brain. 1999;122(9):1613–1628.

82. Mechelli A, Humphreys GW, Mayall K, Olson A, Price CJ. Differential effects of word length and visual contrast in the fusiform and lingual gyri during reading. Proc Biol Sci. 2000;267(1455):1909–1913.

83. Kitada R, Johnsrude IS, Kochiyama T, Lederman SJ. Brain networks involved in haptic and visual identification of facial expressions of emotion: an fMRI study. NeuroImage. 2010;49(2):1677–1689.

84. Daneault V, Hébert M, Albouy G, et al. Aging reduces the stimulating effect of blue light on cognitive brain functions. Sleep. 2014;37(1):85–96.

85. Lok R, Smolders K, Beersma DGM, de Kort YAW. Light, alertness, and alerting effects of white light: a literature overview. J Biol Rhythms. 2018;33(6):589–601.

86. Forkel SJ, Thiebaut de Schotten M, Kawadler JM, Dell’Acqua F, Danek A, Catani M. The anatomy of fronto-occipital connections from early blunt dissections to contemporary tractography. Cortex. 2014;56:73–84.

87. Pantazatos SP, Yanagihara TK, Zhang X, Meitzler T, Hirsch J. Frontal-occipital connectivity during visual search. Brain Connect. 2012;2(3):164–175.

88. Herbet G, Zemmoura I, Duffau H. Functional anatomy of the inferior longitudinal fasciculus: from historical reports to current hypotheses. Front Neuroanat. 2018;12:77.

89. Hepworth LR, Rowe FJ, Walker MF, et al. Post-stroke visual impairment: a systematic literature review of types and recovery of visual conditions. Ophthalmol Res Int J. 2016;5:1–43.

90. Schakel W, Bode C, Elsman EBM, et al. The association between visual impairment and fatigue: a systematic review and meta-analysis of observational studies. Ophthalmic Physiol Opt. 2019;39(6):399–413.

91. Nisenbaum R, Reyes M, Mawle A, Reeves W. Factor analysis of unexplained severe fatigue and interrelated symptoms: overlap with criteria for chronic fatigue syndrome. Am J Epidemiol. 1998;148(1):72–77.

92. Potaznick W, Kozol N. Ocular manifestations of chronic fatigue and immune dysfunction syndrome. Optom Vis Sci. 1992;69(10):811–814.

93. Badham SP, Hutchinson CV. Characterising eye movement dysfunction in myalgic encephalomyelitis/chronic fatigue syndrome. Graefe’s Arch Clin Exp Ophthalmol. 2013;251(12):2769–2776.

94. Becker K, Kohen R, Lee R, et al. Poststroke fatigue: hints to a biological mechanism. J Stroke Cerebrovasc Dis. 2015;24(3):618–621.

95. Lassen NA, Ingvar DH, Skinhøj E. Brain function and blood flow. Scientific Am. 1978;239(4):62–71.

96. Grajny K, Pyata H, Spiegel K, et al. Depression symptoms in chronic left hemisphere stroke are related to dorsolateral prefrontal cortex damage. J Neuropsychiatry Clin Neurosci. 2016;28(4):292–298.

97. Grieve SM, Korgaonkar MS, Koslow SH, Gordon E, Williams LM. Widespread reductions in gray matter volume in depression. NeuroImage: Clin. 2013;3:332–339.

98. Egorova N, Cumming T, Shirbin C, Veldsman M, Werden E, Brodtmann A. Lower cognitive control network connectivity in stroke participants with depressive features. Transl Psychiatry. 2018;7(11):4.

99. Sheline YI, Price JL, Yan Z, Mintun MA. Resting-state functional MRI in depression unmasks increased connectivity between networks via the dorsal nexus. Proc Natl Acad Sci U S A. 2010;107(24):11020–11025.

100. Kaiser RH, Andrews-Hanna JR, Wager TD, Pizzagalli DA. Large-scale network dysfunction in major depressive disorder: a meta-analysis of resting-state functional connectivity. JAMA Psychiatry. 2015;72(6):603–611.

101. Hwang JW, Egorova N, Yang XQ, et al. Subthreshold depression is associated with impaired resting-state functional connectivity of the cognitive control network. Transl Psychiatry. 2015;5(11):e683–e683.

102. Liu J, Ren L, Womer FY, et al. Alterations in amplitude of low frequency fluctuation in treatment-naïve major depressive disorder measured with resting-state fMRI. Hum Brain Mapp. 2014;35(10):4979–4988.

103. Egorova N, Veldsman M, Cumming T, Brodtmann A. Fractional amplitude of low-frequency fluctuations (fALFF) in post-stroke depression. NeuroImage: Clin. 2017;16:116–124.

104. Wang L, Dai W, Su Y, et al. Amplitude of low-frequency oscillations in first-episode, treatment-naive patients with major depressive disorder: a resting-state functional MRI study. PLoS One. 2012;7(10):e48658.

105. Wu S, Mead G, Macleod M, Chalder T. Model of understanding fatigue after stroke. Stroke. 2015;46(3):893–898.

106. Wechsler D. Wechsler Adult Intelligence Scale—Third edition. The Psychological Corporation; 1997.

107. Brandt J. The hopkins verbal learning test: development of a new memory test with six equivalent forms. Clin Neuropsychol. 1991;5(2):125–142.

108. Osterrieth PA. Le test de copie d’une figure complexe. Contribution à l’étude de la perception et de la mémoire. Arch Psychol (Geneve) 1944;30:206–353.

