## Supplemental Material for "Post-stroke fatigue is linked to resting state posterior hypoactivity and prefrontal hyperactivity"

#### Supplementary materials

List of abbreviations

Supplementary tables I-VIII

Supplementary figures I-III

### List of abbreviations

| | |
|---|---|
| **PSF** | Post-stroke fatigue |
| **QoL** | Quality of life |
| **OCSP** | Oxfordshire Community Stroke Project |
| **POCI** | Posterior circulation infarction |
| **mRS** | Modified Rankin scale |
| **RAS** | Reticular activating system |
| **fMRI** | Functional magnetic resonance imaging |
| **BOLD** | Blood-oxygen level dependent |
| **FST** | Fronto-striatal-thalamic |
| **MS** | Multiple sclerosis |
| **PFC** | Prefrontal cortex |
| **GPA** | Granulomatosis with polyangiitis |
| **TBI** | Traumatic brain injury |
| **CFS** | Chronic fatigue syndrome |
| **rs-fMRI** | Resting-state functional magnetic resonance imaging |
| **ALFF** | Amplitude of low-frequency fluctuations |
| **fALFF** | Fractional amplitude of low-frequency fluctuations |
| **CANVAS** | Cognition and neocortical volume after stroke study |
| **PHQ-9** | Patient Health Questionnaire |
| **DSM-V** | Diagnostic and Statistical Manual of Mental Disorders |
| **NIHSS** | National Institutes of Health Stroke Scale |
| **GAD-7** | Generalised Anxiety Disorder questionnaire |
| **T2DM** | Type 2 diabetes mellitus |
| **HVLT-R** | Hopkins verbal learning test revised |
| **ROCF** | Rey-Osterrieth complex figure task |
| **MP-RAGE** | Magnetization-prepared rapid acquisition with gradient echo |
| **TR** | Repetition time |
| **TE** | Echo time |
| **SPM** | Statistical Parametric Mapping |
| **MCAR** | Missing completely at random |
| **mPFC** | Medial prefrontal cortex |

| | |
|---|---|
| **dlPFC** | Dorsolateral prefrontal cortex |

### Supplementary tables

#### Supplementary Table I

*Effect Size Statistics and Interpretations Used for Analyses*

| Effect Size | Notation | Analysis | Interpretation |
|---|---|---|---|
| Cohen's delta | *d* | Independent samples *t*-test | Small = .20, Medium = .50, Large = .80[a] |
| Correlation coefficient | *r* | Mann-Whitney *U* test | Small = .10, Medium = .30, Large = .50[a, b] |
| Cramer's *V* | *V* | Pearson's chi-square test<br>Fisher's exact test | Small = .10, Medium = .30, Large = .50[a, c] |

[a] Cohen (1988). [b] Absolute values of the correlation coefficient will be reported to aid interpretation as per Cohen (1988). [c] Interpretation for 1 degree of freedom

**Supplementary Table II**

*Results for Shapiro-Wilks Test of Normality*

| Variable | Fatigue Status | *W* | *p*-Value (2-tailed) |
|---|---|---|---|
| PHQ-9 | Non-fatigued | .81 | .006 |
| | Fatigued | .84 | <.001 |
| Lesion volume $mm^3$ | Non-fatigued | .70 | <.001 |
| | Fatigued | .66 | <.001 |
| NIHSS Baseline | Non-fatigued | .91 | .150 |
| | Fatigued | .89 | .004 |
| Education | Non-fatigued | .95 | .482 |
| | Fatigued | .90 | .009 |
| Age | Non-fatigued | .86 | .022 |
| | Fatigued | .96 | .217 |
| GAD-7 Score | Non-fatigued | .73 | .001 |
| | Fatigued | .71 | <.001 |
| WAIS-III Digit Span *z*-score | Non-fatigued | .91 | .113 |
| | Fatigued | .96 | .371 |
| HVLT-R *z*-score | Non-fatigued | .95 | .449 |
| | Fatigued | .97 | .467 |
| REY Delay *z*-score | Non-fatigued | .99 | .999 |
| | Fatigued | .96 | .237 |
| Cogstate™ detection *z*-score | Non-fatigued | .96 | .750 |
| | Fatigued | .95 | .113 |
| Cogstate™ identification *z*-score | Non-fatigued | .93 | .303 |
| | Fatigued | .89 | .005 |
| Cogstate™ one-back *z*-score | Non-fatigued | .95 | .594 |
| | Fatigued | .90 | .009 |

*Note*. *N* non-fatigued = 18, *N* fatigued = 45. Degrees of freedom for non-fatigued = 15. Degrees of freedom for fatigued = 31. *W* = *W*-statistic computed from Shapiro-Wilk test; PHQ-9 = Patient Health Questionnaire[48]; NIHSS = National Institutes of Health Stroke Scale[52]; GAD-7 = Generalised Anxiety Disorder questionnaire[54]; WAIS-III = Wechsler Adult Intelligence Scale 3rd Edition[106]; HVLT = Hopkins Verbal Learning Test Revised[107]; ROCF delay = Rey-Osterrieth Complex Figure Delay task[108].

**Supplementary Table III**

*Results for Levene's Test for Equality of Variances for Independent Samples* t*-test*

| Variable | $F$ | $p$-Value (2-tailed) |
|---|---|---|
| PHQ-9 | 5.73 | .020 |
| Lesion volume $mm^3$ | 0.06 | .814 |
| NIHSS Baseline | 0.33 | .569 |
| Education | 1.61 | .210 |
| Age | 0.47 | .498 |
| GAD-7 Score | 1.53 | .221 |
| WAIS-III Digit Span $z$-score | $3.06 \times 10^{-5}$ | .996 |
| HVLT-R $z$-score | 0.30 | .585 |
| REY Delay $z$-score | 0.09 | .760 |
| Cogstate™ detection $z$-score | 3.02 | .088 |
| Cogstate™ identification $z$-score | 0.16 | .694 |
| Cogstate™ one-back $z$-score | 1.33 | .253 |

*Note*. Degrees of freedom = 1. $F$ = $F$-statistic computed from Levene's test; PHQ-9 = Patient Health Questionnaire[48]; NIHSS = National Institutes of Health Stroke Scale[52]; GAD-7 = Generalised Anxiety Disorder questionnaire[54]; WAIS-III = Wechsler Adult Intelligence Scale 3rd Edition[106]; HVLT = Hopkins Verbal Learning Test Revised[107]; ROCF delay = Rey-Osterrieth Complex Figure Delay task[108].

**Supplementary Table IV**

*Output for Independent Samples* t*-tests*

| Variables | Non-fatigued | | Fatigued | | *t*(df) | *p* | *d* |
|---|---|---|---|---|---|---|---|
| | *M* | *SD* | *M* | *SD* | | (2-tailed) | |
| WAIS-III Digit span *z*-score | -0.13 | 0.82 | -0.03 | 0.90 | -0.40 (59) | .691 | .11 |
| HVLT-R *z*-score | 0.28 | 1.18 | 0.09 | 1.10 | 0.60 (61) | .550 | .17 |
| ROCF Delay *z*-score | 0.004 | 1.20 | -0.16 | 1.22 | 0.47 (59) | .641 | .13 |
| Cogstate™ detection *z*-score | 0.19 | 0.31 | -0.15 | 0.55 | 2.46 (60) | .017 | .76 |

*Note*. WAIS-III = Wechsler Adult Intelligence Scale 3rd Edition[106]; HVLT-R = Hopkins Verbal Learning Test Revised[107]; ROCF delay = Rey-Osterrieth Complex Figure Delay task[108]; *d* = Cohen's delta statistic.

**Supplementary Table V**

*Output for Chi-Square and Fisher's Exact Tests*

| Variables | Non-fatigued | | Fatigued | | $\chi^2(df)$ | *p* (2-tailed) | *V* |
|---|---|---|---|---|---|---|---|
| | *N* | % | *N* | % | | | |
| *N* female | 5 | 27.78 | 17 | 37.78 | 0.57 (1) | .564 | .10 |
| *N* with depression [a] | 0 | 0.00 | 19 | 42.22 | | .001 | .42 |
| *N* with mRS ≥ 2 [a] | 4 | 22.22 | 15 | 33.33 | | .545 | .12 |
| Stroke side [a] | | | | | | | |
| *N* Left | 8 | 44.44 | 17 | 37.78 | | | |
| *N* Right | 9 | 50.00 | 27 | 60.00 | | .520 | .11 |
| *N* Bilateral | 1 | 5.56 | 1 | 2.22 | | | |
| Subtype of Ischaemic stroke | | | | | | | |
| *N* LACI | 4 | 22.22 | 6 | 13.33 | | | |
| *N* PACI | 9 | 50.00 | 26 | 57.78 | 0.79 (2) | .686 | .11 |
| *N* POCI | 5 | 27.78 | 13 | 28.89 | | | |
| *N* TACI | 0 | 0.00 | 0 | 0.00 | | | |
| *N* with T2DM at baseline [a] | 0 | 0.00 | 12 | 26.67 | | .013 | .31 |
| *N* with ischaemic heart disease at baseline | 1 | 5.56 | 4 | 8.89 | 0.20 (1) | .999 | .06 |
| *N* with hypertension at baseline | 9 | 50.00 | 27 | 60.00 | 0.53 (1) | .576 | .09 |

*Note*. *V* = Cramer's *V* statistic; mRS = Modified Rankin Scale[53]; LACI = lacunar cerebral infarction; PACI = partial anterior cerebral infarction; POCI = posterior cerebral infarction; TACI = total anterior cerebral infarction.

[a] Analysed via Fisher's exact test and thus a chi-square statistic is not computed/reported.

**Supplementary Table VI**

*Output for Mann-Whitney* U *Tests*

| Variables | Non-fatigued | | Fatigued | | *U* | *Z* | *p* | *r* |
|---|---|---|---|---|---|---|---|---|
| | *Mdn* | IQR | *Mdn* | IQR | | | | |
| Age | 67.5 | 16 | 71 | 19 | 329.5 | -1.15 | .254 | .14 |
| PHQ-9 | 1 | 2 | 4 | 6 | 134 | -4.16 | <.001 | .52 |
| NIHSS at baseline | 2 | 3 | 2 | 3 | 372.5 | -0.51 | .619 | .06 |
| GAD-7 | 0 | 2 | 3 | 3 | 242.5 | -2.52 | .011 | .32 |
| Education at baseline | 12.5 | 4 | 11 | 5 | 352 | -0.81 | .422 | .10 |
| Lesion volume (ml) | 1.90 | 11.93 | 1.40 | 6.83 | 212 | -0.64 | .534 | .09 |
| Cogstate™ identification *z*-score | 0.05 | 0.70 | -0.12 | 0.60 | 377 | -0.30 | .773 | .04 |
| Cogstate™ one-back *z*-score | -0.57 | 1.45 | -0.57 | 1.73 | 385.5 | -0.16 | .874 | .02 |

*Note*. PHQ-9 = Patient Health Questionnaire[48]; NIHSS = National Institutes of Health Stroke Scale[52]; GAD-7 = Generalised Anxiety Disorder questionnaire[54]; *Mdn* = median; *IQR* = interquartile range; *U* = *U*-statistic from Mann-Whitney *U* test; *Z* = standardised *U* statistic; *r* = correlation coefficient.

**Supplementary Table VII**

*z-fALFF Results for Fatigued vs Non-Fatigued Analysis in the 0.01-0.08 Hz Band Excluding the Participant with Lesion Overlap*

| Region | Hemisphere | x | y | z | Peak *p*(unc) | Cluster *p*(FDR-corr) | *k* | *T* | *Z* |
|---|---|---|---|---|---|---|---|---|---|
| *z*-fALFF 0.01–0.08 non-fatigued > fatigued patients (age, sex, NIHSS) | | | | | | | | | |
| calcarine cortex | bilateral | 6 | -84 | -3 | <.001 | .023 | 53 | 4.22 | 3.92 |
| | | -3 | -84 | -3 | <.001 | | | 4.06 | 3.79 |
| | | 3 | -75 | 9 | <.001 | | | 3.47 | 3.29 |
| lingual gyrus | left | -9 | -72 | 3 | <.001 | .185 | 23 | 4.36 | 4.03 |
| *z*-fALFF 0.01–0.08 fatigued > fatigued patients (age, sex, NIHSS) | | | | | | | | | |
| mPFC | bilateral | -6 | 66 | 9 | <.001 | .219 | 25 | 3.65 | 3.44 |
| | | 0 | 66 | 18 | <.001 | | | 3.64 | 3.44 |

*Note*. *z*-fALFF = *z*-transformed values of fractional amplitude of low-frequency fluctuations; unc = uncorrected; FDR-corr = false discovery rate corrected; *k* = size of cluster in voxels; *T* = *t*-statistic; *Z* = *z*-transformed *t*-statistic; mPFC = medial prefrontal cortex; NIHSS = National Institutes of Health Stroke Scale[52]; x, y, z = coordinates in MNI space (mm).

**Supplementary Table VIII**

*z-fALFF Results for Depressed Fatigued vs Non-Depressed Fatigued Analysis in the 0.01 – 0.08 Hz Band*

| Region | Hemisphere | x | y | z | Peak *p*(unc) | Cluster *p*(FDR-corr) | *k* | *T* | *Z* |
|---|---|---|---|---|---|---|---|---|---|
| z-fALFF 0.01–0.08 depressed > non-depressed patients (age, sex, NIHSS) | | | | | | | | | |
| dlPFC | Left | -36 | 21 | 45 | <.001 | .019 | 57 | 4.46 | 3.99 |
| | | -21 | 30 | 57 | <.001 | | | 4.28 | 3.86 |
| | | -33 | 33 | 48 | <.001 | | | 4.18 | 3.78 |

*Note*. *z*-fALFF = *z*-transformed values of fractional amplitude of low-frequency fluctuations; unc = uncorrected; FDR-corr = false discovery rate corrected; *k* = size of cluster in voxels; *T* = magnitude of *t*-statistic; *Z* = *z*-transformed *t*-statistic; mPFC = medial prefrontal cortex; NIHSS = National Institutes of Health Stroke Scale[52]; x, y, z = coordinates in MNI space (mm).

#### Supplementary figures

##### Supplementary Figure I

*One Participant's Lesion Overlap with Significant Posterior Clusters from Fatigued vs Non-Fatigued 0.01 – 0.08 Hz and Slow-4 Analyses. Participant lesion is shown in green and significant posterior clusters from fatigued vs non-fatigued analysis in the 0.01 – 0.08 Hz band and slow-4 band (0.027 – 0.073 Hz) are shown in red. Note the clear overlap of the lesion and the calcarine cluster.*

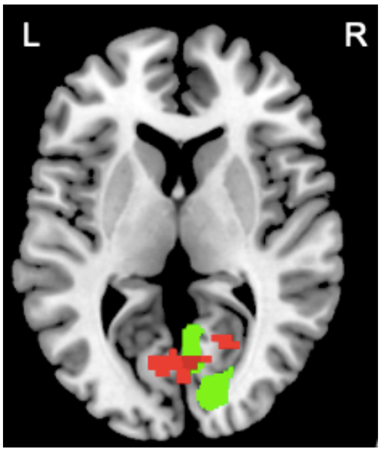

**Supplementary Figure II**

*Main Results for Depressed Fatigued vs Non-Depressed Fatigued Analysis in the 0.01 – 0.08 Hz Band. Results for depressed fatigued > non-depressed fatigue contrast. The colour bar represents the magnitude of the t-statistic computed from the analysis.*

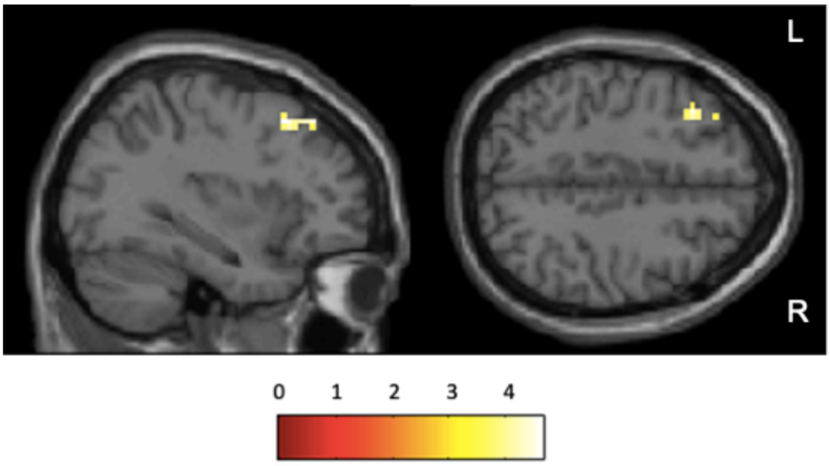

**Supplementary Figure III**

*Overlay of Significant Clusters for Main Analysis (Fatigued vs Non-Fatigued) and Secondary Analysis (Depressed Fatigued vs Non-Depressed Fatigued). Results of the main group comparison (fatigued vs non-fatigued) for all frequencies are shown in red. Results of the secondary group comparison (depressed fatigued vs non-depressed fatigued) for 0.01 – 0.08 Hz are shown in blue. The absence of an overlap suggests that although fatigue in the current study is measured as part of the PHQ-9 (depression) questionnaire, the brain correlates attributable to fatigue and depression can be dissociated.*

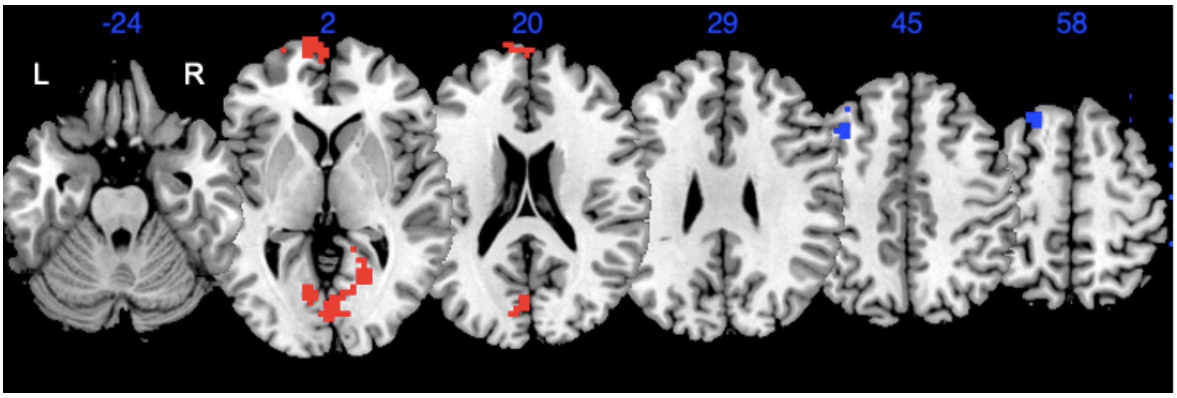
